# Pathways from inequality to health: Social determinants of health shape the gut microbiome in low-resource U.S. communities

**DOI:** 10.64898/2026.07.20.26358501

**Authors:** C Chaney, J Ong, L Kwak, K Grathwohl, S Wang, K Betts, JA Beauregard, KL Nemeth, S Waimon, A Zhang, AB Shing, A Samsonov, M Pfaff-Nash, Braveen Ragunanthan, SS Urlacher, TJ Cepon-Robins, TE Gildner, EK Mallott

**Author notes:** Corresponding authors: Carlye Chaney and Elizabeth Mallott.

## Abstract

The gut microbiome influences cardiovascular and gastrointestinal disease risk, yet the indirect pathways through which structural social determinants of health (SDoH) shape microbial composition remain understudied. We investigated causal pathways linking SDoH—systemic conditions that shape health opportunities—to the gut microbiome in two low-resource, flood-prone U.S. communities. We collected questionnaires, dried blood spots, and fecal samples (n=131; ages 3–79) in Mississippi and Illinois (2022–2023) as part of a longitudinal project on infrastructure and health, identifying bacterial composition via 16S rRNA sequencing. Controlling for covariates, structural equation modeling showed education positively predicted microbiome richness and evenness, partially mediated through income and diet. This suggests that structural inequities in education, food access, and income significantly influence the microbiome. Notably, contrary to established literature, greater consumption of processed and fried foods was associated with increased alpha diversity. Therefore, in this context, diet may covary with environmental vulnerability, such as opportunistic pathogen exposure from frequent flooding. Analysis of beta diversity showed direct effects from education, diet, and CRP; income and homeownership had indirect effects mediated through diet. The taxa driving these relationships likely include Tannerellaceae for diet and a subgroup of Firmicutes for income, based on our differential abundance analysis. These findings indicate that pathways linking SDoH to the microbiome may vary by local infrastructure and environmental context. This research highlights how structural inequality may be biologically embedded, suggesting that public health interventions should target infrastructural inequities linked with environmental racism, rather than individual behavioral change.

## Introduction

Social determinants of health (SDoH) – the structural, upstream forces that influence individual health outcomes regardless of personal behavior or lifestyle choices – are not equally experienced across populations, contributing significantly to racial health disparities in the United States (1,2). SDoH are also associated with alterations in the human gut microbial community, driven by factors such as socioeconomic status, educational attainment, diet quality, social interactions, pollutant exposure, and psychosocial stressors (3–16). Consequently, the human gut microbiome may function as a dynamic system that directly mediates how structural inequality shapes human biology and health. However, our mechanistic understanding of the relationships between the gut microbiome and individual structural determinants remains incomplete. Furthermore, while SDoH appear to exhibit joint effects on the gut microbiome during early life (9), the degree to which these complex, multivariate relationships persist across the adult life course remains unexamined.

Rather than disentangling the role of multiple structural factors, such as SDoH, previous gut microbiome research has often focused on individual factors or used race as a proxy for biological differences to explain the relationship between racial health disparities and alterations to the gut microbiome (17,18). However, race, a social construct, does not have an underlying biological basis (19–24). Instead, structural discrimination often leads people of color to experience greater adverse SDoH, which may exacerbate racial health inequities (25–30). A more precise potential mechanism for the relationship between SDoH and changes in the gut microbiome appears to be inflammation. For example, SDoH like socioeconomic status, neighborhood deprivation, and racial discrimination are associated with elevated levels of several markers of inflammation, including C-reactive protein (CRP) and allostatic load (31–37). Intestinal and systemic inflammation are also robustly associated with altered gut microbiome diversity and composition (11,38,39).

To clarify the relationships between SDoH and the gut microbiome, we examined the effects of multiple SDoH and assessed the mediating effect of inflammation using structural equation modeling (SEM) with data from the Rural Embodiment and Community Health (REACH) Study (reachresearch.org). The REACH Study uses detailed surveys alongside biosample collection — including dried blood spots and fecal samples — to examine how environmental conditions (particularly repeated flooding and water quality concerns) affect human well-being and health outcomes. The communities participating in the study are two low-resource, majority-Black communities in the United States located in southwestern Illinois and the Delta region of Mississippi, both of which face chronic flooding and deteriorating water and sewer infrastructure (40–44). Preliminary research from the Mississippi site indicated high rates of intestinal parasite infection among children who live in an area that experiences flooding from the local bayou, and this parasite infection was significantly associated with elevated intestinal inflammation (40,41). These results suggest that structural factors may influence environmental exposures in these contexts, including exposure to pathogens spread through contaminated water and soil, with downstream effects on numerous aspects of human biology and health, including potential links to the gut microbiome. Therefore, collecting data from these two sites enhances our understanding of the gut microbiome in two key ways. First, we investigate how a range of environmental conditions and lived experiences shapes gut microbiota diversity and composition in the United States (Box 1) by testing a theoretical model of the ways that SDoH are related to each other and the gut microbiome. Second, the majority of microbiome data lacks racial, ethnic, economic, and ecological diversity, and most research in the U.S. focuses on higher-income, white, urban populations (45–47). This study reduces these biases in microbiome data by recruiting individuals in low-resource, majority-Black communities. Currently We used this unique dataset to 1) identify which SDoH variables are associated most strongly with the gut microbiome in low resource communities, including which variables have direct and/or indirect effects; and 2) examine if inflammation, measured using C-reactive protein and an allostatic load index, mediates the relationship between SDoH and the gut microbiome. This research advances our understanding of the pathway through which multiple SDoH are physiologically embedded without using race as a proxy for structural conditions, further elucidating the relationship between SDoH, inflammation, and the gut microbiome.

### Box 1.

**Summary alpha and beta diversity measures used in this study.**

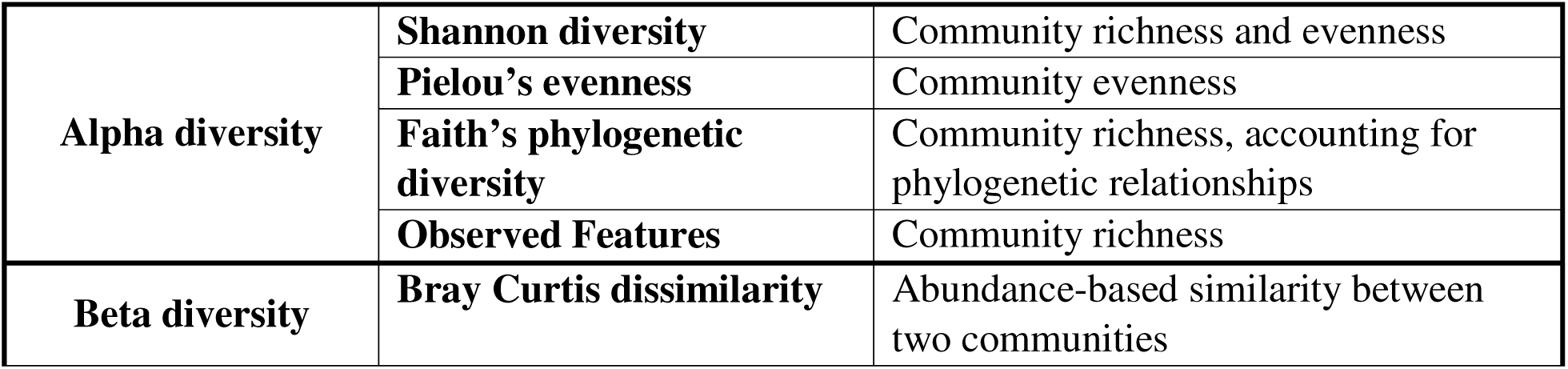
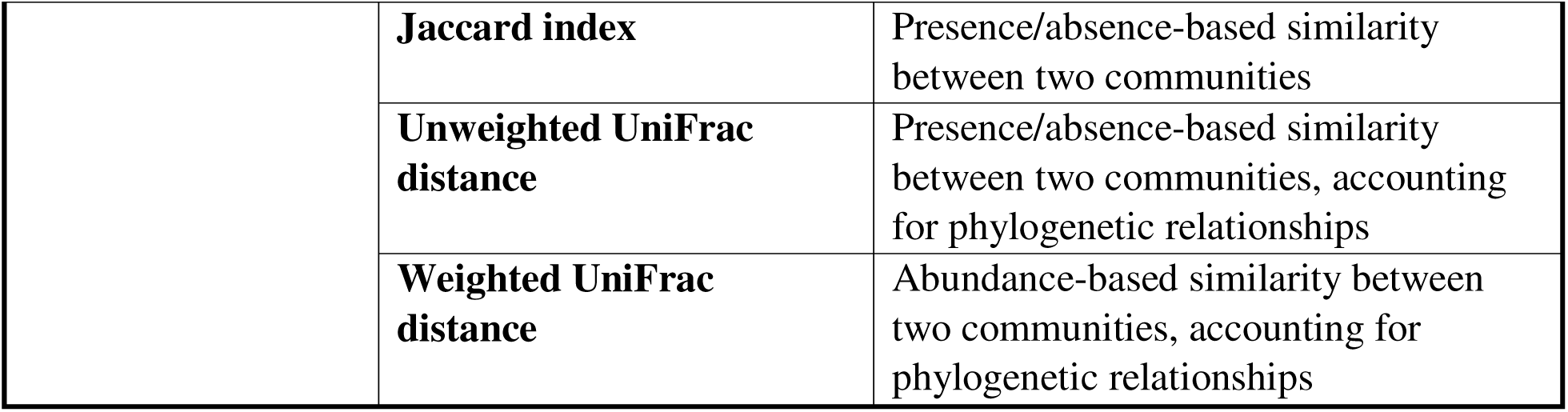

## Results

### Study Population

Participants were recruited from two study sites (Mississippi Delta and southwestern Illinois) in 2022 and 2023 as part of the REACH Study – a longitudinal project investigating the relationships between environmental factors - such as severe flooding and poor water infrastructure - living conditions, and health inequities. As previously described (48), the MS study site is located in the Mississippi Delta region with a population of approximately 2,000 individuals, of which around 95% identify as Black or African American. The median household income is $20,265 based on US census estimates (49). The second study site is located in southwestern Illinois with a population of roughly 14,500 people. Within this study site, roughly 65% of individuals identify as Black or African American based on US census data and the household median income is $30,000 (50,51). The final combined sample size was 131 individuals, which included 79 participants from IL and 52 from MS. Participants were roughly evenly divided between study years (IL 2022 = 38 individuals, MS 2022 = 26; IL 2023 = 41, MS 2023 = 26). Descriptive statistics and summary information for the sample are provided in Tables 1 and 2, respectively. Descriptive statistics for each study site are included in Supplementary Table 1. The distribution of participants across racial and employment categories was significantly different by study site (p<0.05), so we controlled for study site in all models. Specifically, IL had a greater percentage of white participants compared to MS (16.5% vs 1.9%). IL also had a greater number of participants who worked part-time or were unemployed while MS had a greater proportion of participants who were unable to work or whose job did not fit the other employment categories.

**Table 1:** Descriptive statistics for continuous variables, mean and standard deviation (SD); n = 131. Dairy consumption is an additive score (range: 0-8) based on frequency of consumption of dairy products. Fatty food consumption is an additive score (range 0-27) based on consumption of seven fatty foods (fast food, fried food, processed meat, high fat salad dressings, processed snacks, high fat cooking oils, and red meat).

| Variable | Mean (SD) | Range |
| --- | --- | --- |
| Age, years* | 45.5 (22.4) | 3-79 |
| BMI, kg/m<sup>2</sup> | 31.5 (8.7) | 14.2-67.9 |
| CRP, mg/L | 4.4 (6.3) | 0.02-44.2 |
| Allostatic load score** | 3.3 (1.1) | 1-6 |
| Dairy consumption | 5.5 (1.8) | 2-8 |
| Fatty food consumption | 17.8 (4.3) | 8-27 |

\* Distribution of children's ages is provided in the Supplementary Material
\*\*Due to missing data, the sample size for analyses including the allostatic load score was 101 individuals (missing data was distributed randomly between IL and MS).

**Table 2:** Summary information for categorical variables, number of individuals (%); n = 131.

| Variable | Number of Individuals (%) |
| --- | --- |
| Sex |  |
| Female | 92 (70.2%) |
| Male | 39 (29.8%) |
| Race* |  |
| Black/African American | 104 (79.4%) |
| White | 14 (10.7%) |
| Multiracial or Other | 13 (9.9%) |
| Education± |  |
| Less than high school | 12 (9.2%) |
| High school | 50 (38.2%) |
| College, no degree | 41 (31.3%) |
| Associate | 12 (9.2%) |
| Bachelor's | 12 (9.2%) |
| Master's | 4 (3.1%) |
| Employment±* |  |
| Full-time | 27 (20.6%) |
| Part-time | 21 (16.0%) |
| Unemployed | 25 (19.1%) |
| Retired | 33 (25.2%) |
| Unable to work (disabled) | 17 (13.0%) |
| Other | 8 (6.1%) |
| Healthcare access** |  |
| Sufficient | 20 (15.3%) |
| Limited | 111 (84.7%) |
| Food security |  |
| Did not skip meals | 94 (71.8%) |
| Skipped meals regularly during a 1–2-month period | 17 (13.0%) |
| Skipped meals regularly during a 3–6-month period | 12 (9.2%) |
| Skipped meals regularly for 7+ months | 8 (6.1%) |
| Household overcrowding*** | 20 (15.3%) |
| Household income range |  |
| <\$10,000 | 32 (24.4%) |
| \$10,000-19,999 | 36 (27.5%) |
| \$20,000-34,999 | 31 (23.7%) |
| \$35,000-49,999 | 14 (10.7%) |
| \$50,000-74,999 | 14 (10.7%) |
| \$75,000+ | 4 (3.1%) |

± Data for adults only
\* Significantly different by site ( $p < 0.05$ )
\*\* Healthcare access was assessed by asking about medical source for regular doctor appointments, where primary care was coded as 1 (sufficient) and going to either the emergency room or urgent care was coded as 2 (insufficient).
\*\*\* Defined as more than 1.01 person per room (excluding bathrooms and kitchen) as per the U.S. Census Bureau

### Income, diet quality, education, and homeownership predict alpha diversity

The alpha diversity SEM models (Shannon diversity, Pielou’s evenness, Faith’s PD, and Observed Features; see Box 1) based on our preregistered conceptual diagram did not fit the data well based on Fisher’s C statistic. Fisher’s C tests the null hypothesis that the predicted model and observed data are equal; a non-significant result indicates good model fit. Initial models used either CRP modeled continuously or allostatic load as a measure of inflammation. Diet was measured using either PCA scores derived from an unhealthy food survey or frequency scores for sugar and dairy consumption. In analyses, we used PCA1, which accounted for 26% of dietary variation; the largest loadings were for fried food, processed food, and baked goods. Therefore, we used tests of directed separation to identify strong relationships between variables that were not included in the original model (see Supplementary Material). We refined our model by including these relationships, which improved both Fisher’s C and Akaike Information Criterion (AIC). We present the results from the refined model below.

Income, diet, and education significantly predicted alpha diversity in models that used PCA dietary scores and CRP as the measure of inflammation (we present CRP results as they had the larger sample size; allostatic load results were generally similar and can be found in the Supplementary Material). Income and diet demonstrated consistent direct effects: income significantly predicted Shannon diversity (*β* = 0.22; bias-corrected and accelerated confidence interval (CI) = 0.11 to 0.38), Pielou’s evenness (*β* = 0.19; CI = 0.07 to 0.33), and observed features (*β* = 0.19; CI = 0.05 to 0.36), while diet significantly predicted Shannon diversity (*β* = 0.16; CI = 0.02 to 0.30) and evenness (*β* = 0.15; CI = 0.01 to 0.28). Several variables showed significant indirect effects. Education indirectly influenced Shannon diversity (*β* = 0.05; CI = 0.02 to 0.11), evenness (*β* = 0.04; CI = 0.01 to 0.1), and observed features (*β* = 0.05; CI = 0.01 to 0.12). Home ownership exerted a negative indirect effect on Shannon diversity (*β* = −0.03; CI = −0.092 to −0.003). In addition to its direct effects, income had a negative indirect effect on evenness (*β* = −0.07; CI= −0.14 to −0.01). The models fit the data well for the SEM models investigating Shannon Diversity (Fisher’s C = 92.78, p = 0.88), Pielou’s Evenness (Fisher’s C = 93.55, p = 0.87), Faith’s PD (Fisher’s C = 93.52, p = 0.87), and observed features (Fisher’s C = 91.53, p = 0.90). Significant results for models predicting Shannon diversity are depicted in Fig 1A. Detailed results for all variables, including the direct and indirect effects, as well as the results from the models using sugar and dairy frequency scores are provided as tables in the Supplementary Material.

**Figure 1:**
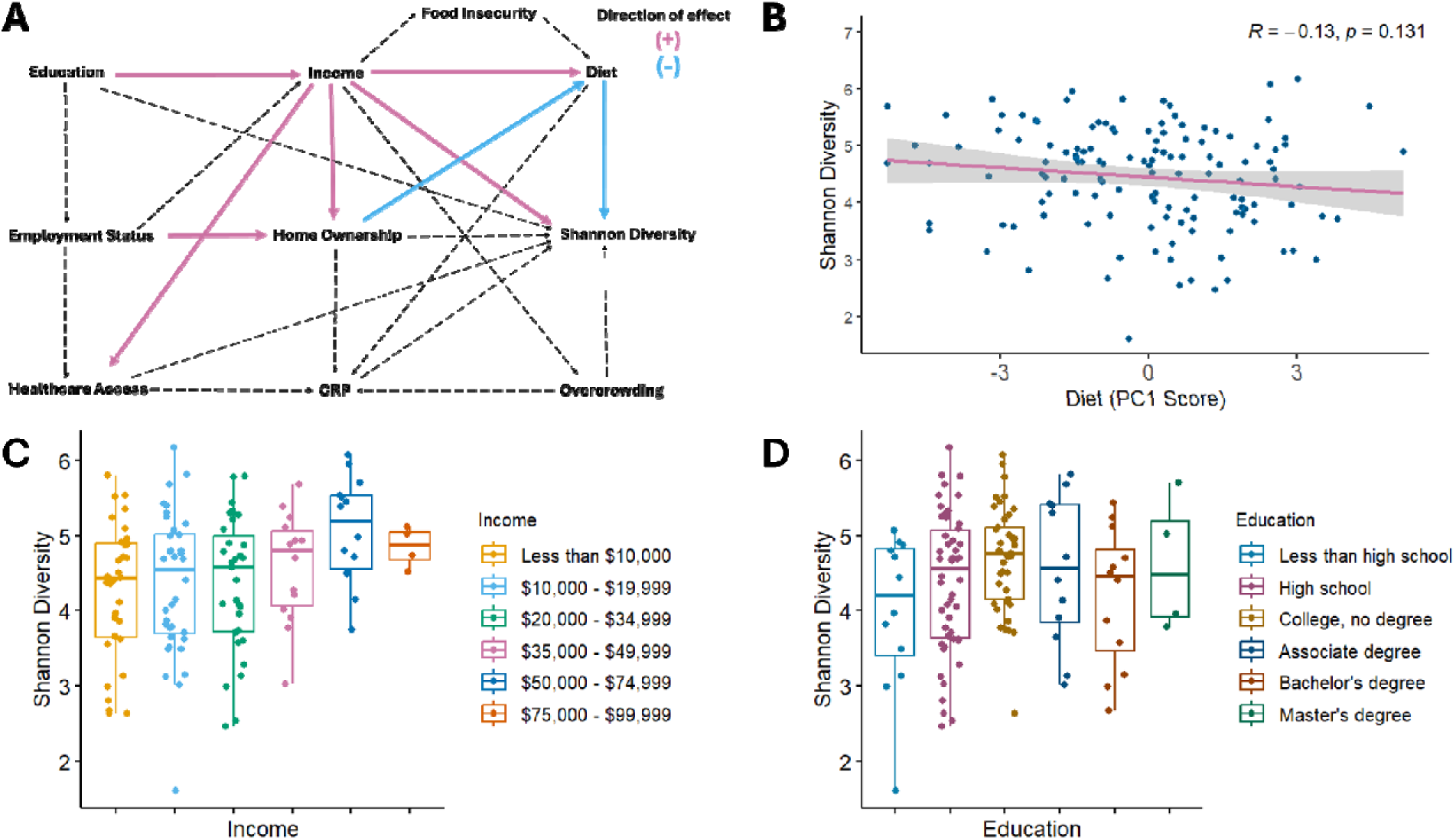
Shannon diversity results A) Piecewise SEM results for the model predicting Shannon Diversity using CRP as a measure of inflammation and diet measured from unhealthy food consumption scores (pink = significant positive relationships, blue = significant negative relationship). B) Correlation plot of Shannon diversity index and diet quality based on PC1 scores. C) Boxplot of Shannon diversity index by income range. D) Boxplot of Shannon diversity index by education level. Diet quality in 1A and 1B have been reverse-coded for ease of visual interpretation so that greater dietary quality scores indicate greater healthy food consumption.

The relationship between inflammation and alpha diversity metrics was similar in models with allostatic load instead of CRP, which also fit the data well based on Fisher’s C (Shannon diversity: Fisher’s C = 94.12, p = 0.89; Pielou’s Evenness: Fisher’s C = 94.41, p = 0.88; Faith’s PD: Fisher’s C = 91.92, p = 0.92; observed features: Fisher’s C = 91.94, p = 0.92). Detailed results for all variables, as well as the results from the models using sugar and dairy frequency scores, are described in the Supplementary Material.

### Inflammation, education, diet quality, homeownership, income, and overcrowding predict variation in microbial community composition

Similar to the alpha diversity models, our initial SEM models for beta diversity (see Box 1) initially showed poor fit. We subsequently used tests of directed separation to identify missing relationships. In the final models utilizing PCA dietary scores and CRP, education, diet, and CRP emerged as significant predictors. Education, diet, and CRP demonstrated consistent direct effects on beta diversity. Education significantly predicted Bray Curtis NMDS1 (*β* = −0.15; CI = −0.28 to −0.02), Jaccard index NMDS1 (*β* = −0.14; CI = −0.27 to −0.02), and unweighted UniFrac NMDS1 (*β* = −0.15; CI = −0.29 to −0.03). Diet had direct effects on Bray Curtis NMDS1 (*β* = −0.16; CI = −0.32 to −0.03), Jaccard index NMDS1 (*β* = −0.24; CI = −0.37 to −0.12), and weighted UniFrac NMDS1 (*β* = 0.16; CI = 0.02 to 0.30). Additionally, CRP exerted a direct effect on Bray Curtis NMDS3 (*β* = −0.18; CI = −0.35 to −0.03). Several variables influenced beta diversity through indirect pathways. Home ownership had indirect effects on the Jaccard index NMDS1 (*β* = 0.05; CI = 0.01 to 0.11) and weighted UniFrac NMDS1 (*β* = −0.03; CI = −0.09 to −0.01). Income also indirectly influenced Jaccard index NMDS1 (*β* = 0.07; CI = 0.01 to 0.13) and weighted UniFrac NMDS1 (*β* = −0.08; CI = −0.16 to −0.02). Finally, overcrowding exerted indirect effects on Bray Curtis NMDS3 (*β* = 0.03; CI = 0.01 to 0.08) and weighted UniFrac NMDS2 (*β* = −0.02; CI = −0.05 to −0.001). Following the inclusion of d-separation paths, the models demonstrated strong fit for Bray Curtis (Fisher’s C = 97.73, p = 0.96), Jaccard (Fisher’s C = 112.69, p = 0.76), unweighted UniFrac (Fisher’s C = 102.11, p = 0.93), and weighted UniFrac (Fisher’s C = 96.43, p = 0.91). Statistically significant relationships for the Bray Curtis dissimilarity model are depicted in Fig 2A. Full results for all variables and models using sugar and dairy frequency scores are provided in the Supplementary Material.

**Figure 2:**
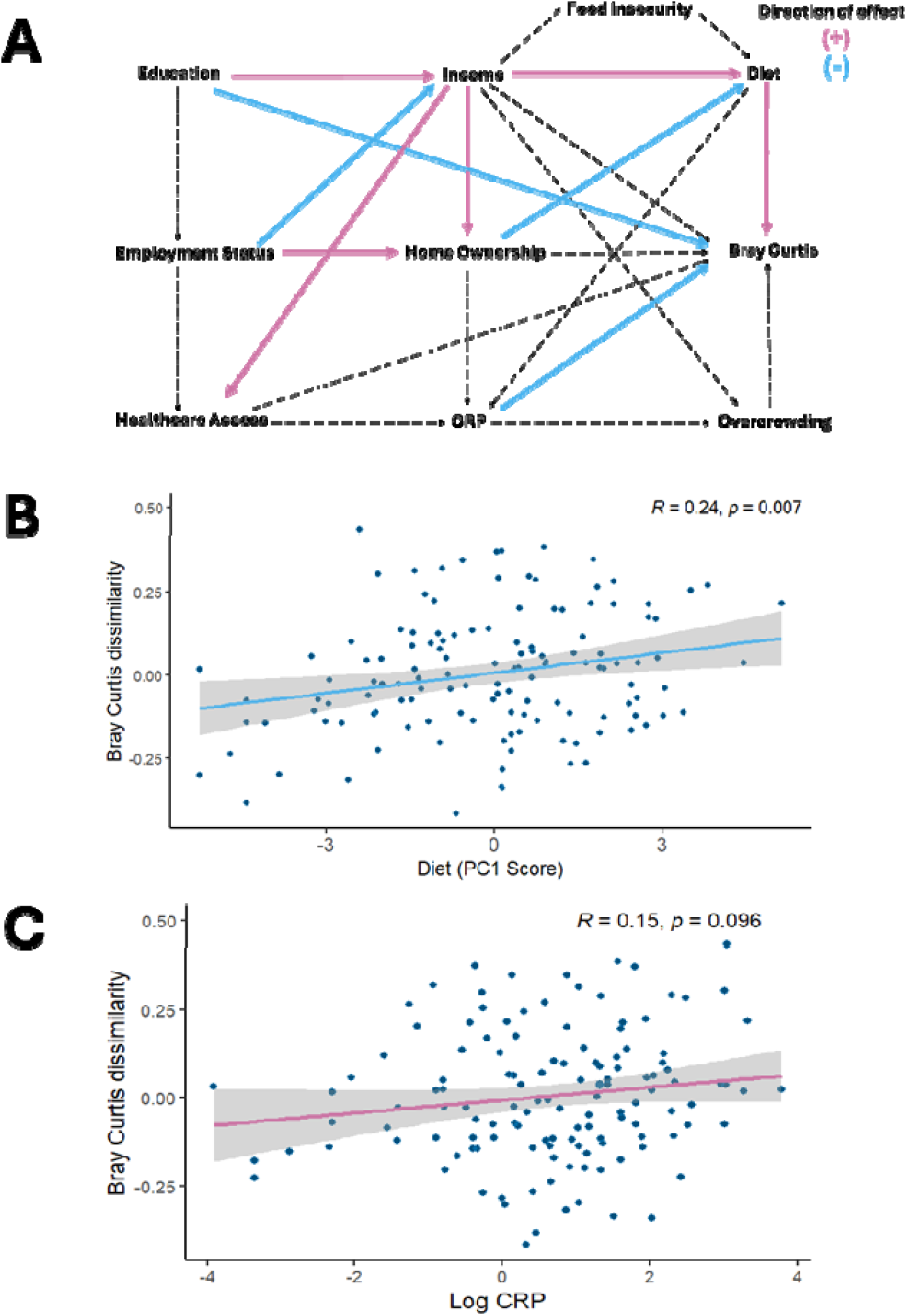
Bray Curtis dissimilarity results. A) Piecewise SEM results for the model predicting Bray Curtis dissimilarity using CRP as a measure of inflammation and diet measured from unhealthy food consumption scores (pink = significant positive relationships, blue = significant negative relationship). B) Correlation plot of Bray Curtis dissimilarity and diet quality based on PC1 scores. C) Correlation plot of Bray Curtis dissimilarity and log C-reactive protein (CRP) levels. Diet quality in 2A and 2B have been reverse-coded for ease of visual interpretation so that greater dietary quality scores indicate greater healthy food consumption.

When accounting for allostatic load, patterns for education, diet (PC1), and allostatic load trended similarly to the CRP models; however, within these models, only diet remained a significant direct predictor. Several lifestyle and health factors demonstrated direct effects on beta diversity. Diet directly predicted the Jaccard index NMDS1 (*β* = −0.22; CI = −0.41 to −0.07). Healthcare access predicted both Jaccard index NMDS2 (*β* = −0.21; CI = −0.41 to −0.02) and weighted UniFrac NMDS2 (*β* = 0.20; CI = 0.01 to 0.37). As healthcare access was coded as 1 for sufficient and 2 for insufficient, the negative coefficient for Jaccard NMDS2 indicates that scores increase with greater healthcare access. Home ownership showed a negative association with Jaccard index NMDS2 scores (*β* = −0.18; CI = −0.37 to 0.00). Smoking significantly predicted Bray Curtis MDS1 (*β* = −0.16; CI = −0.36 to −0.02), while BMI predicted weighted UniFrac NMDS1 (*β* = −0.22; CI = −0.41 to −0.03). Employment was the only variable with indirect effects, and it was associated with both Jaccard index NMDS2 (*β* = −0.07; CI = −0.16 to 0.00) and weighted UniFrac NMDS2 (*β* = 0.07; CI = 0.02 to 0.15). The data fit the models well for all metrics (Bray Curtis: Fisher’s C = 95.35, p = 0.95; Jaccard: Fisher’s C = 100.43, p = 0.90; Unweighted UniFrac: Fisher’s C = 99.23, p = 0.92; Weighted UniFrac: Fisher’s C = 91.56, p = 0.92). Full results for all variables, including models utilizing sugar and dairy frequency scores, are available in the Supplementary Material.

### Income and relative abundance of microbial taxa

We then investigated if shifts in the relative abundance of microbial taxa may drive our alpha and beta diversity results using ALDEX2 (52,53). The relative abundance of most taxa was not associated with the SDoH variables used in this study. At the phyla level, we found that the highest income group had significantly lower levels of Firmicutes D (estimate = −3.83, adjusted p-value = 0.01). We also found marginally significant differences associated with PC1 scores for Tannerellaceae (Bacteroidota) (estimate = −0.73, adjusted p-value = 0.08).

**Figure 3:**
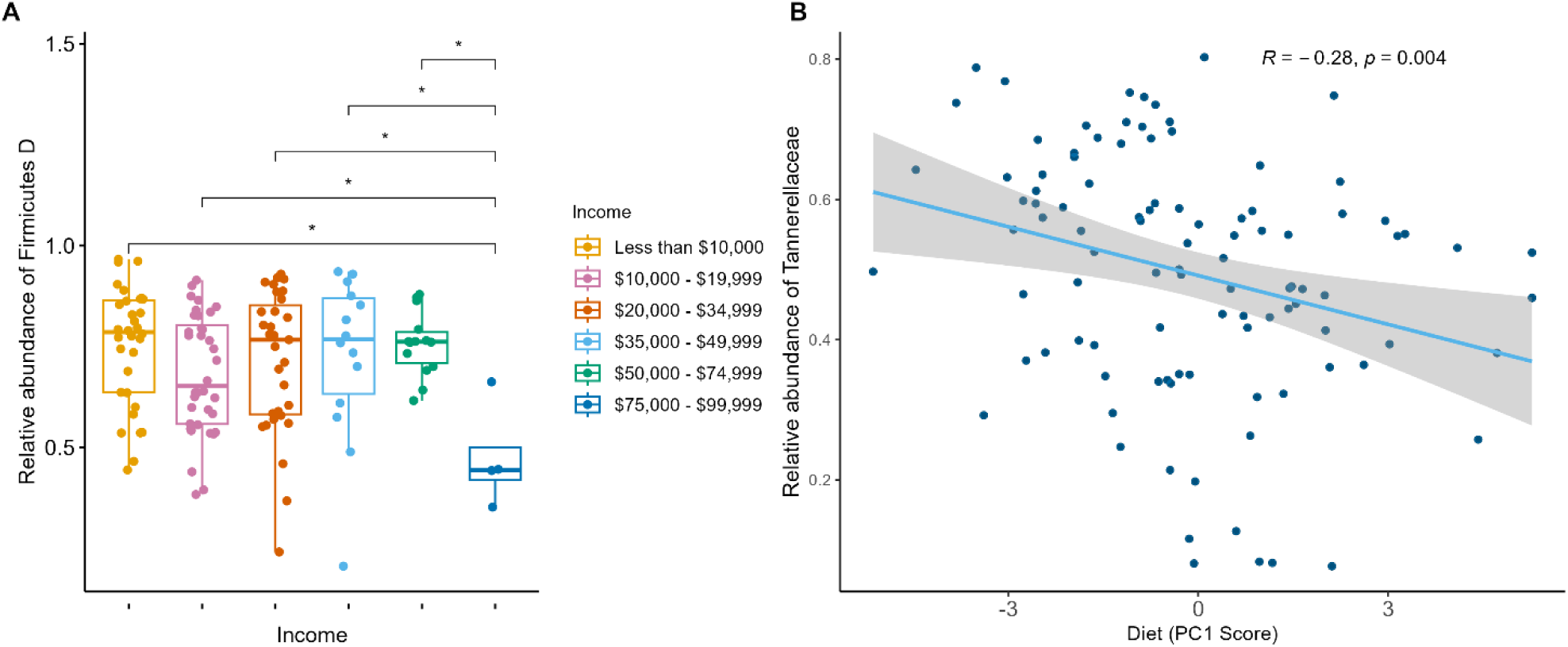
Relative abundance results A) Boxplot of the relative abundance of Firmicutes D by income range. B) Correlation plot of relative abundance of Tannerellaceae (Bacteroidota) by diet quality as measured by PC1 diet scores.

## Discussion

In this study, a piecewise SEM approach identified pathways linking environmental conditions to microbiome diversity and composition in two low-resource U.S. communities with frequent and severe flooding, suggesting that SDoH are physiologically embedded via changes in the gut microbiome. Our results demonstrate how piecewise SEMs can be a valuable tool for understanding the physiological embedding of the structural environment when sample sizes are small. Specifically, we found that education had indirect effects on both alpha and beta diversity. The positive effect of education on alpha diversity was mediated through greater income and diet, suggesting that improvements in education access may have downstream benefits on income, diet, and alpha diversity. In beta diversity models, education, diet, and CRP showed direct effects while income and homeownership had indirect effects that were mediated through diet, indicating that education and diet are also important drivers of gut microbiome composition. The taxa contributing to these relationships may include a subgroup of Firmicutes for income and Tannerllaceae for diet based on our differential abundance analysis. Additionally, our model did not replicate previously identified relationships between several SDOH variables, such as the relationship between income and food security, indicating that the pathway linking SDoH variables may vary based local contexts.

Income and diet were both directly, positively associated with multiple measures of alpha diversity. Income had the largest effect size of all direct predictors and was associated with increased alpha diversity – a metric associated with improved health outcomes (54–58). This result echoes those of prior research using multivariate modeling approaches (3,7,12), and likely represents individuals’ greater ability to purchase healthy foods, better access to health resources, and greater access to greenspaces. However, diet showed unexpected results; we found that greater consumption of fried food, processed food, and baked goods was associated with greater alpha diversity – an effect opposite of those previously established patterns in the literature, where consumption of processed foods is generally inversely associated with alpha diversity (59–66). Therefore, the dietary survey may have functioned as a proxy for an underlying, unmeasured factor with direct effects on alpha diversity. Rather than indicating a healthy gut, this elevated diversity may stem from an environmental influx of opportunistic, soil- and water-borne pathogens introduced during frequent municipal infrastructure failures, such as the chronic flooding and frequent sewage backups in these communities (40–44). In this context, unhealthy food consumption likely serves as a behavioral marker of environmental vulnerability and the chronic stress associated with such infrastructure failures. More research is needed to investigate this potential pathway.

Diet had the opposite effect when analyzing its role as a mediator via indirect effects: we identified a novel pathway from income to microbiome diversity, mediated by diet. Specifically, higher income was associated with lower consumption of fatty, sugary, and dairy products, which in turn correlated with greater alpha diversity. This finding responds to the call in the literature for research quantifying both the direct and indirect relationships between socioeconomic status, diet, and gut microbiome changes (67). Our approach extends previous work by identifying the indirect effects of education that were mediated through income and diet. Taken together, this pathway suggests that improvements in education access in these communities may drive downstream improvements in the mediators of income and healthy food access, which could then increase alpha diversity and, by extension, reduce chronic disease risk and improve health outcomes including cardiovascular disease, allergies, irritable bowel syndrome, anxiety, and stress (54–58).

While the beta diversity results varied across the metrics we used, we found that inflammation, education, diet, homeownership, income, and overcrowding were associated with gut microbiome composition. Yet, the differences across SEM models using different beta diversity metrics provide detail on the varied ways that SDoH may be physiologically embedded through microbiome composition. For example, variation in low abundance or rare taxa was explained by the indirect effect of education and direct effects of income and healthcare access on Jaccard distances and unweighted UniFrac distances (see Box 1). However, differences in rare and potentially closely related taxa were better explained by the indirect effect of employment status, which was significant for Jaccard distances but not unweighted UniFrac. This relationship was mediated through homeownership and diet. Lastly, adiposity may be influencing the abundance of common, closely related taxa through inflammation, as BMI had an indirect effect on Bray-Curtis dissimilarity mediated through CRP. By comparing differences in patterns across these beta diversity models, a SEM approach reveals fine-grained patterns in the ways that different SDoH interact to influence aspects of microbiome composition.

The taxa underlying the relationships between SDoH and alpha or beta diversity may include the Firmicutes D phyla and Tannerellaceae family. Specifically, income was negatively associated with the abundance of Firmicutes D, while Tannerellaceae was positively associated with diet quality. Firmicutes D includes all members of the class Bacilli, which is a class of generally Gram-positive, spore-forming, aerobic bacteria, although some are facultative anaerobes (68,69). While many Bacilli are nonpathogenic, a few can cause disease, such as *Bacillus anthracis* (anthrax) and *Bacillus cereus* (food poisoning), so higher levels of this class among lower-income individuals may suggest that they are at greater risk for these pathogens. Additionally, although some *Bacilli* species may have some capacity for butyrate synthesis (70,71), the class is not considered to be a major butyrate producer (72,73). Therefore, individuals with greater levels do not benefit from the multiple positive benefits of microbial short-chain fatty acid production, including greater protection against inflammation (74). This taxonomic shift provides an example of how environmental racism is biologically embedded. Members of the Bacilli class are robust, spore-forming environmental bacteria prevalent in contaminated soils and stagnant, post-flood waters (75,76). Lower-income individuals in these regions face disproportionate exposure to pathogens due to structural housing inequities and proximity to overflowing, underfunded wastewater systems.

For diet, greater consumption of unhealthy food was marginally associated with lower levels of Tannerellaceae. Several species in this family are positively related to human health, such as *Parabacteroides distasonis and P. goldseinii*, which may decrease inflammation, obesity risk, and risk of rheumatoid arthritis, in addition to benefiting metabolic function (77,78). However, these effects may be context dependent, as *P*. *distasonis* also appears to have adverse effects in immunocompromised individuals and may contribute to autoimmune disorders, such as Type 1 diabetes (79–81). We lack the sample size to detect effects at lower levels of taxonomic classification, which would have allowed us to better understand which species contributed to the observed differences in microbial diversity and composition.

The pathways identified in our SEM models also failed to reproduce several well-supported population-level patterns, indicating the importance of local context for physiological embedding. For example, in our refined models, we did not observe a relationship between income and food security (82–88). However, in our study sites, people frequently report utilizing food pantries (unpublished data). As our food insecurity question focused on skipping meals due to a lack of food, these food pantries may provide a local safety net that disconnects the expected relationship between income and food security, as measured by skipping meals because of financial concerns. This decoupling highlights a localized resilience strategy against systemic economic disenfranchisement, where mutual aid networks and food pantries act as vital safety nets. However, while these pantries mitigate caloric scarcity, they are frequently constrained to distributing shelf-stable, highly processed items, limiting the quality of accessible food. We also failed to observe a relationship between education and employment, which is possibly due to the relatively older age of our sample (i.e., 35% of participants in our sample were 60 years old or older). These results highlight the importance of considering local context when investigating the relationships between SDoH, biomarkers, the gut microbiome, and health outcomes, as the local environment may produce distinct pathways or fail to replicate well-established pathways observed at the population level.

Our unexpected findings, such as those related to inflammation and food security, may be due to relevant study limitations. First, we were unable to include measures of the built environment, such as flooding exposure or frequency of sewage backups into the home, which are common in these study sites (43), and our models indicated that much variation in the gut microbiome remains unexplained. Second, despite robust research demonstrating the contributions of inflammation to gut dysbiosis (11,38,39), we did not find robust support for systemic inflammation as the biological mechanism linking SDoH and alpha diversity using CRP, and we found no support for allostatic load mediating relationships between SDoH and gut microbiome diversity or composition. As CRP and allostatic load measure whole-body inflammation, a more local measure of intestinal-specific inflammation might have provided more robust results and additional biomarkers could provide a more nuanced understanding of inflammatory processes, such as interleukin-6, interleukin-10, erythrocyte sedimentation rate, or procalcitonin. Lastly, our food insecurity question only captured issues related to scarcity, not quality of food, and future work in these communities is needed to disentangle different aspects of food security and its relationship with gut microbiome diversity and composition. These three main limitations provide opportunities for future research to better understand the physiological embedding of SDoH and further characterize local versus population-level patterns. Working in collaboration with local communities to address their research priorities, the REACH Study has future analyses planned to address several of these knowledge gaps. Specifically, ongoing and future work will incorporate geospatial household flood exposure data, environmental microbiome analyses, additional health outcomes, and contaminant exposure to further characterize the ways the environmental racism is embedded in these contexts.

## Conclusions

In this study, we identified a causal pathway linking multiple adverse SDoH to changes in gut microbiome diversity and composition in two low resource U.S. communities, demonstrating one way in which structural inequality can become physiologically embedded. Based on effect sizes, reductions in poverty through structural interventions to reduce pay inequity or increase economic opportunities would likely provide the greatest improvements in microbiome richness and evenness with downstream benefits for chronic disease risk. Additionally, education and diet showed robust effects across several beta diversity metrics, suggesting that interventions to increase educational opportunities and access to fresh, nutrient-rich foods would have the broadest benefits to the gut microbiome. Contrary to our predictions, inflammation was only associated with one beta diversity metric, Bray-Curtis. More research is needed to understand what other physiological mechanisms link SDoH to changes in the gut microbiome. The changes in alpha and beta diversity related to income may be driven by differences in abundance of a subgroup of Firmicutes (Firmicutes D) that includes many pathogenic species that provide little-to-no anti-inflammatory benefits. Additionally, greater consumption of fatty, fried, and fast food could lead to changes in the gut microbiome through altered abundance of Tannerellaceae, which may have negative health effects in immunocompromised individuals and contribute to the development of autoimmunity.

However, further work is needed to clarify the relationships of these taxa with health outcomes. Larger sample sizes and longitudinal data are needed to identify other taxa driving these relationships. Our SEM models also revealed that well-established relationships between SDoH variables may not apply in some low resource contexts; for example, we did not find a relationship between income and food insecurity, potentially because of the easy access to and lack of stigmatization associated with food pantries in these study sites. However, this food may not provide high quality nutrition, and more research is needed to understand the nuances of this finding. Additionally, some of our unexpected findings, such as the counter-intuitive relationship between diet and microbiome richness, may be explained by aspects of the local built environment that were not included in the present analyses, such as deteriorating and inadequate water and sewage infrastructure. Taken together, these results demonstrate how confirmatory path analysis can be used to better understand the relationships between different aspects of the environmental conditions and the physiological embedding of structural inequality through changes in gut microbiome diversity and composition while highlighting several avenues for further research.

## Materials and Methods

### Participant recruitment and data collection

Participants were recruited in the summer of 2022 and 2023 in both rural Mississippi and southwestern Illinois; participants received a gift card and a subset of their individual health results. Study team members interviewed participants using a questionnaire covering the domains of demographics, household characteristics, health, behavior, activities, and diet. As part of demographic data collection, we used US census defined categories for race. Overcrowding was defined as more than 1.01 person per room (excluding bathrooms and kitchen) (89). The diet survey asked participants to report the weekly frequency of consuming 16 food and beverage groups (coffee/caffeinated drink, alcoholic drinks, candy, baked goods, frozen desserts, carbonated drinks, fast food, fried food, red meat, processed meat, butter/margarine, milk products, eggs, high fat dressings, processed snacks) (see Supplementary Material). Healthcare access was assessed by asking about medical source for regular doctor appointments, where primary care was coded as 1 (sufficient) and going to either the emergency room or urgent care was coded as 2 (insufficient). Food insecurity was assessed using a question that is part of the USDA food insecurity questionnaire (90); the question was, “In the last 12 months, did you or other adults in the household ever cut the size of your meals or skip meals because there wasn’t enough money for food?” and answer options included never, one-two months, three-six months, or seven or more months. Additionally, anthropometric measurements (height, weight, body fat percentage, blood pressure) were collected.

We also collected dried blood spots (DBS) to measure CRP, an indicator of systemic inflammation. The DBS were collected using standard protocols (91). We then dried the filter paper at room temperature for 4 hours before freezing it at −20°C. Samples were then shipped on dry ice to the Urlacher Human Evolutionary Biology and Health Lab (HEBHL) at Baylor University for long term storage at −80°C and biomarker analysis. Additional measures of cholesterol, triglycerides, white blood cell count, and hemoglobin were collected from the same finger-prick samples using point-of-care devices (Cardiochek Plus Analyzer, Hemocue WBC System, Hemocue HB 201 System).

Lastly, following previously published protocols (40), participants self-collected stool samples using stool specimen collection pans. Samples were kept at ambient temperature and returned to the study team within 24 hours. After collection, samples were aliquoted into sterile 2ml cryovials and frozen at −80C until analysis in the Mallott Laboratory. In total, 251 fecal samples were obtained.

### CRP Analysis

The DBS were analyzed for CRP concentrations in the HEBHL using a modified high-sensitivity enzyme-linked immunosorbent assay (ELISA) protocol optimized for DBS (92–94). All samples were run in duplicate, with controls on each plate. Mean intra-assay and inter-assay *CV*s were 2.87% and 1.68%, respectively. Measured CRP values falling below the detection limit of the assay (*n* = 9) were assigned a value of the lower limit of detection divided by the square root of two. To enhance interpretability, CRP DBS concentrations were converted to serum-equivalent values using a validated conversion equation (94,95).

### Allostatic Load

We created an allostatic load score to measure chronic inflammatory processes based on the specific cardiovascular, metabolic, and immune biomarkers available in the REACH Study dataset. Allostatic load represents the cumulative physiological dysregulations due to repeated responses to extreme conditions, like chronic stress (96). Due to its effects on the neuroendocrine, immune, metabolic and cardiovascular systems (97), it is a measure that includes at least one biomarker of immune, metabolic, and cardiovascular function. Biomarker values are indexed based on clinical values to determine individual risk of disease or poor health. Common biomarkers used for allostatic load calculation are summarized in Duong et al. (2017) (98). In our study, we created our allostatic load index from blood pressure, hemoglobin levels, non-HDL cholesterol levels, BMI, triglyceride levels, CRP concentrations, and white blood cell counts. The cutoffs for elevated risk for each clinical marker were defined as the following: blood pressure risk was systolic blood pressure above 129 or diastolic blood pressure above 80 based on American Heart Association guidelines or healthy blood pressure while taking medicine to treat high blood pressure (99); CRP risk was defined as greater than 3 mg/L (95,100); hemoglobin risk was less than 12 g/dL for women and less than 13 g/dL for men (101); triglyceride risk was defined as greater than 150 mg/dL based on CDC guidelines or healthy triglyceride levels while taking a medication for high cholesterol (102); non-HDL risk was defined as greater than 130 mg/dL or healthy non-HDL levels while taking a medication for high cholesterol (103); BMI risk was defined as greater than 30 kg/m² or less than 18.5 kg/m² based on CDC guidelines for healthy weight (104); and white blood cell count was defined as less than 5 mg/dL or greater than 10 mg/dL for men and less than 4.5 mg/dL and greater than 11 mg/dL for women based on guidelines by the Leukemia and Lymphoma Society (105).

To determine this subset of immune, metabolic, and cardiovascular markers, we used data from the 2005/2006 and 2017/2018 waves of National Health and Nutrition Survey (NHANES data). Specifically, we used raw NHANES data (n=5913) to analyze different subsets of biomarkers representative of allostatic load and compared their performance to previously published allostatic load methods in the NHANES dataset (106,107). In models to assess the fit of our allostatic load measure, we found that our measure explained less variance (Adj R2 = 0.05) compared to previous measures (Adj R2 = 0.17) in models containing the poverty-to-income ratio, gender, age, smoking, education, and ethnicity; however, the patterns of the results fit predictions for allostatic load better. Specifically, allostatic load was higher among men in our model and white individuals had the lowest allostatic load; allostatic load was also associated with greater age in both metrics, as expected (108,109). In the REACH variables, we did not find significant associations between smoking and income-to-poverty ratio using the NHANES data. When we plotted ethnicity*gender using the NHANES data, the REACH allostatic load measure found that white women had the lowest allostatic load and Black women the highest, which fits expected patterns based on previous research (109,110) and is likely due to differences in exposure to chronic discrimination and other SDoH. The previous allostatic load measures used in Chaney & Wiley (2023) and Shirazi et al. (2020) also fit this pattern, but the fit was not as strong (106,107). Therefore, we concluded that our allostatic load measure was sufficiently comparable to other published methods.

### DNA Extractions

DNA was extracted from 243 samples using the Powerlyzer Powersoil DNA Isolation Kit. The manufacturer’s protocols were followed with the modification of a 10-minute incubation at 65 C after C1 solution is added (40,41). We included extraction negatives with no stool sample with each extraction batch to control for contamination.

### 16S rRNA gene amplification, sequencing, and analysis

After DNA extractions, a 16S rRNA gene amplification was performed using the 515F and 926R primer set using established protocols (111,112). If samples did not initially amplify, 0.75 μL of dimethyl sulfoxide (DMSO) was added to each polymerase chain reaction (PCR), reducing the H_2_O by an equal volume. We performed a PCR negative was included on each plate where molecular grade water was used instead of sample DNA to control for contamination. Samples were sequenced at the Genome Technology Access Center at the McDonnell Genome Institute at Washington University in St. Louis. Samples were sequenced on an Illumina MiSeq (2x300bp) platform.

### Data Analysis

Our research question, conceptual model, and analysis plans were preregistered on Open Science Framework (https://osf.io/cd4gk/), and code for all analysis can be found at https://github.com/Mallott-Lab/REACH_SDoH. Data supporting the findings of this study are available from the NCBI Database of Genotypes and Phenotypes (dbGaP) at [Insert Link/Accession Number]. Access is restricted to authorized researchers upon approval of a Data Use Certification.

We created two separate measures of diet quality. First, we ran principal components analysis on all diet quality questions. While 80% of the variance was represented by 8 principal components (PC), we chose to analyze only PC1, which contained the greatest amount of variance (26%), as including 8 PCs inflated the AICs of our models and led to overfitting issues. Second, we created additive scores for consumption of fatty foods, sugary foods, and dairy given their known impacts on the human gut microbiome (59, 61–66, 113).

Fatty food was represented by frequency of consuming fast food, fried food, processed meat, high fat salad dressings, processed snacks, high fat cooking oils, and red meat. Sugary food consumption was calculated based on frequency of consuming candy, baked goods, and frozen desserts. Dairy consumption was based on frequency of consuming butter or margarine and milk products. As fatty food consumption and sugary food consumption were moderately correlated (0.44), we dropped sugary food consumption from our models as it was more highly correlated with dairy consumption than fatty food consumption and only investigated fatty food and dairy consumption.

Raw 16S amplicon sequence reads obtained were demultiplexed, paired, and denoised using the Divisive Amplicon Denoising Algorithm pipeline (DADA2) in QIIME2 v2024.2 (114,115). We exported sequences to R (v4.2.3) to run SCRuB (Source-tracking for Contamination Removal in microBiomes), a method to detect and remove contamination by incorporating information on samples and controls (116,117). Decontaminated ASVs were exported back into QIIME2 for further processing. We aligned the ASVs with MAFFT and constructed a phylogeny with FastTree (Katoh et al., 2002; Price et al., 2010). After rarefaction, a sample size of 176 individuals remained. We then calculated four measures of alpha diversity: Shannon diversity, Pielou’s evenness, Faith’s Phylogenetic Diversity, and observed features. We also calculated four measures of beta diversity: Jaccard Index, Bray-Curtis dissimilarity, and unweighted and weighted UniFrac distances. We used Non-metric Multidimensional Scaling (NMDS) to cluster groups of samples based on k-means and chose the k parameter that resulted in the lowest stress value (k = 3 for Jaccard Index, Bray Curtis dissimilarity, and unweighted UniFrac; k = 2 for weighted UniFrac).

Downstream statistical analyses and visualizations were carried out in R v4.2.3 (R Core Team, 2023). We used pairwise structural equation modelling (*PSEM* package) to assess our conceptual model (118). Within the *PSEM* package, we used tests of directed separation to identify potentially missing relationships in order to refine the model and assessed model fit based on Fisher’s C and Akaike Information Criterion. We performed differential abundance analysis to investigate income, PC1 scores, and healthcare access. We did not find any significant taxa using ANCOM-BC (119) or a GLMM with a negative binomial distribution; as the false discovery rate adjusted p-values were all 1, it is likely that our sample size was too small to detect effects with these methods. We then used ALDEX2 as research suggests it performs better than other differential abundance methods with small sample sizes (52,53). All models took into account various covariates that could be confounding our analysis: age, BMI, site, study year, smoking status, and antibiotics use in the last month. For individuals who participated in both study years (n=5), we dropped their data from the first study year. After this step, 131 individuals remained with complete data for our variables of interest.

### Conceptual model validation with NHANES data

To validate our conceptual model, we used NHANES data. NHANES is a national biannual cross-sectional survey that collects information on health and nutrition from a nationally representative sample of children and adults living in the United States. We used measurements from the 2017-2018 data. As the *PSEM* package cannot hand sample weights and strata and we did not need the analysis to be generalizable to the US population, we did not incorporate sample weights and strata. We cleaned data and selected variables to parallel those available through the REACH Study, and we built the SEM according to our original conceptual model. As noted above, we used tests of directed separation to identify potentially missing relationships in order to refine the model and assessed model fit based on Fisher’s C and Akaike Information Criterion. See Supplementary Material for conceptual model validation results using this data.

## Supporting information

Supplementary Model Results

## Data Availability

All data produced in the present study are available upon reasonable request to the authors.

## Acknowledgments

We would like to thank all study participants and community partners, including the Centreville Citizens for Change, Nicole Nelson, Kalila Jackson, Maliaka Hill, Kennedy Gardner, AN, JC, and CF. We also acknowledge funding from NSF SPRF #2313258, NSF GCR #2523966, Washington University in St. Louis Seeding Projects for Enabling Excellence & Distinction (SPEED) Award, and the Transdisciplinary Institute in Applied Data Science (TRIADs) at Washington University in St. Louis.

